# Deriving OCT-Equivalent Retinal Nerve Fiber Layer Thickness Maps from Fundus Photographs with Deep Learning Improves Glaucoma Diagnosis

**DOI:** 10.64898/2026.05.26.26354047

**Authors:** Lily Shi, Min Shi, In Young Chung, Louis R. Pasquale, Lucy Q. Shen, Mengyu Wang

**Affiliations:** Harvard Ophthalmology AI Lab, Schepens Eye Research Institute, Massachusetts Eye and Ear, Harvard Medical School, Boston, MA, USA; School of Computing & Informatics, University of Louisiana at Lafayette, Lafayette, LA, USA; Department of Ophthalmology, Massachusetts Eye and Ear, Harvard Medical School, Boston, MA, USA; Department of Ophthalmology, Icahn School of Medicine at Mount Sinai, New York, NY, USA; Kempner Institute for the Study of Natural and Artificial Intelligence, Harvard University, Boston, MA, USA; Harvard Data Science Initiative, Harvard University, Boston, MA, USA; Broad Institute of MIT and Harvard, Boston, MA, USA

## Abstract

**Purpose:** To develop and evaluate a deep learning model that predicts optical coherence tomography (OCT)–equivalent retinal nerve fiber layer thickness (RNFLT) maps directly from color fundus photographs and to assess their diagnostic value for glaucoma detection.

**Design:** Retrospective model development and evaluation study.

**Participants:** 15,031 paired fundus photographs and spectral-domain OCT scans collected at Massachusetts Eye and Ear between 2011 and 2022.

**Methods:** Paired fundus and OCT images were used to train a U-Net–based model to predict pixel-wise RNFLT maps with artifact-corrected supervision. Diagnostic performance was evaluated across single-modality models (fundus photos only, real RNFLT maps, predicted RNFLT maps) and multimodal fusion models (fundus + predicted RNFLT maps). Stratified analyses examined model performance across glaucoma severity and demographic subgroups. Glaucoma was defined based on standard criteria applied to Humphrey 24-2 visual field testing.

**Main Outcome Measures:** Mean absolute error (MAE) and structural similarity index (SSIM) for RNFLT map prediction. Area under the ROC curve (AUC) and accuracy for glaucoma detection.

**Results:** RNFLT map prediction achieved a MAE = 15.4 μm and a SSIM = 0.65, measured against artifact-corrected RNFLT maps derived from OCT. For glaucoma detection, the predicted RNFLT-only classifier outperformed the fundus-only classifier (AUC 0.889 vs 0.883, p < 0.005; Accuracy 82.0% vs 78.0%), but performed worse than the real-RNFLT-only classifier (AUC 0.889 vs 0.903, p < 0.005). Multimodal fusion of fundus images with predicted RNFLT maps improved performance, achieving an AUC of 0.909, outperforming all single-modality inputs (p < 0.005 vs fundus-only, predicted-RNFLT-only, and real-RNFLT-only). Performance gains between the fundus-only and the multimodal classifier were greater in early-stage glaucoma compared to severe cases: accuracy increased from 55.3% to 64.0% in mild cases, from 71.5% to 80.4% in moderate cases, and from 90.0% to 94.6% in severe cases.

**Conclusions:** Predicted RNFLT maps derived from fundus photographs provide quantitative, OCT-like structural information and improve glaucoma detection. Unlike prior work that predicted only summary RNFLT values, our model generates full RNFLT maps that better support glaucoma classification than fundus images alone. This approach offers a scalable pathway for early glaucoma screening and expands diagnostic access in resource-limited settings.

## Introduction

Glaucoma is an irreversible optic neuropathy and a leading cause of blindness worldwide, projected to affect over 110 million people by 2040.^1^ The disease is characterized by progressive loss of retinal ganglion cells and their axons in the retinal nerve fiber layer (RNFL), leading to visual field deterioration. Because early stages are often asymptomatic, glaucoma often goes undiagnosed until the disease is advanced. It is estimated that approximately 50% of individuals with glaucoma in the United States are unaware of their condition.^2^ Early detection is critical to prevent permanent vision loss, yet widespread screening remains challenging.

Traditionally, clinicians have relied on fundus photography to identify structural changes associated with glaucoma, such as increased optic cup-to-disc ratio and RNFL defects. In recent decades, optical coherence tomography (OCT) has transformed ophthalmic imaging by providing quantitative, depth-resolved measurements of RNFL thickness (RNFLT). OCT is now considered the gold standard for structural glaucoma evaluation in clinical practice.^3^ However, OCT acquisition is technically demanding and operator-dependent, and segmentation errors can introduce artifacts that degrade RNFLT measurements.^4^ In addition, OCT devices are less widely available, particularly in primary care settings where large-scale population screening may be needed and in resource-limited environments, where infrastructure and personnel constraints limit their use.^5^

By contrast, fundus photography is more broadly available.^6^ However, its 2D images offer limited ability to identify subtle or early changes in the optic nerve head and RNFL. Additionally, interpretations of fundus images often depend on subjective evaluations by clinicians, leading to inconsistent results and reducing the reliability of fundus photography for detecting glaucoma, particularly in its early stages.^7–8^ These trade-offs motivate methods that can deliver OCT-like structural information derived from fundus photos. Deep learning has been extensively studied for automated glaucoma detection using retinal imaging. Fundus photograph datasets are far more numerous and larger in volume than OCT datasets, making them more accessible for developing AI models.^9^ Fundus-based models trained on color photographs have achieved high performance with area under the curve (AUC) values above 0.9,^10–12^ yet these results might be affected by label-source bias, as many datasets derive glaucoma labels from clinician interpretation of disc photos rather than objective functional tests or structural criteria, limiting generalizability.^13^ OCT-based models, which quantify structural features such as RNFL thickness or ganglion cell-inner plexiform layer (GCIPL) thickness, provide objective and reproducible imaging biomarkers.^14^ Previously, our team demonstrated that CNNs trained on OCT-derived RNFLT maps outperformed those trained on fundus images in glaucoma diagnosis, confirming that OCT captures measurable structural damage rather than surface-level appearance, making it a more objective and informative basis for diagnosis.^13^

Recent “machine-to-machine” (M2M) approaches have shown that deep networks can infer OCT-based metrics directly from fundus photographs. Medeiros et al. first demonstrated that global mean RNFLT values could be predicted from optic disc photos with high correlation to true OCT measurements, and subsequent studies extended this framework to regional RNFLT values or circumpapillary sectors.^15–18^ These models validated that fundus images contain sufficient information to approximate OCT-based measures of axonal loss in glaucoma. More recently, Liu et al. applied an M2M-trained RNFLT predictor to longitudinal fundus images and showed that the predicted values correlated with glaucoma progression risk.^19^ However, most prior studies predicted global or regional summary values rather than generating full RNFLT maps, limiting their ability to demonstrate the overall and localized structural loss in glaucoma. In contrast, generating full RNFLT maps provides continuous, pixel-level structural information. In addition, the predicted maps mirror the format of OCT output, allowing clinicians to visually assess localized patterns of RNFL loss. The only study on full map synthesis used a small dataset and did not establish downstream diagnostic utility.^20^ To date, no study has demonstrated that synthesized, pixel-wise RNFLT maps meaningfully improve glaucoma diagnostic performance.

In this study, we present an end-to-end framework that (i) predicts pixel-wise OCT-equivalent RNFLT maps directly from color fundus photographs, and (ii) evaluates the clinical utility of these synthesized maps for glaucoma detection.

## Methods

### Dataset

This study utilized a dataset of fundus photographs (Zeiss Visucam, Carl Zeiss AG), spectral-domain OCT scans (Cirrus OCT, Carl Zeiss AG), and standard automated perimetry (visual field, or VF) tests collected at Massachusetts Eye and Ear between 2011 and 2022. This retrospective study was approved by the Mass General Brigham Institutional Review Board (IRB) and conducted in accordance with the principles outlined in the Declaration of Helsinki. The IRB also deemed informed consent to be waived for this study.

A total of 16,936 paired fundus–OCT images were initially identified that met the following criteria to ensure high-quality and temporally aligned data: OCT scans were required to have a signal strength of at least 6 out of 10^13^; VFs were required to meet reliability criteria, defined as fixation losses ≤ 33%, false-negative rates ≤ 20%, and false-positive rates ≤ 20%^13^; Visual field (VF) tests were performed within 30 days of the OCT exam; and fundus photographs were obtained within 180 days of the OCT exam.

For glaucoma labeling, we followed widely used functional criteria^21–22^: eyes were classified as glaucomatous if they had a VF mean deviation (MD) < –3 decibels (dB), an abnormal glaucoma hemifield test (GHT), and an abnormal pattern standard deviation (PSD). Non-glaucomatous eyes had MD ≥ –1 dB, normal GHT, and PSD probability > 0.05.

An additional 1,905 samples were later removed based on RNFLT map quality criteria, as described below, resulting in 15,031 samples used for analysis. Because the dataset included multiple imaging pairs per patient, we split the dataset at the patient level to ensure that no patient’s data appeared in more than one of the training, validation, or test sets, thereby preventing information leakage.

### Deep Learning Modeling Development and Training

To improve the predictive accuracy of RNFLT maps, we used artifact-corrected OCT-derived targets and a weighted loss function that assigned greater influence to high-quality regions during training. We evaluated diagnostic performance across single-modality classifiers (fundus images, real RNFLT maps, predicted RNFLT maps) as well as multimodal fusion architectures, including a dual-branch mid-level fusion and an attention-based fusion model that dynamically weighted cross-modal features. All models were trained and validated on the final dataset (n = 15,031). We additionally performed stratified analyses by glaucoma severity and demographic subgroups (race, ethnicity, sex, and age) to assess model sensitivity to early structural loss and evaluate the consistency of performance across subgroups.

### OCT to RNFLT Map Conversion

RNFLT maps were generated from Cirrus HD-OCT volumetric scans by computing the distance between the inner limiting membrane (ILM) and the RNFL segmentation boundaries. The difference was scaled using the device-specific axial pixel spacing factor of 0.00196 mm/pixel, to convert the values to microns.^13^ Optic disc and cup regions were excluded using segmentation-derived masks and encoded with categorical values to distinguish them from invalid tissue (0=invalid, 1=valid tissue, 2=disc, 3=cup). Invalid tissue referred to pixels where RNFL < ILM, which yielded a negative thickness. These pixels were marked as invalid and set to NaN (Not a Number) so they did not contribute to downstream losses or evaluation metrics. Valid tissue referred to pixels within the analyzable RNFL region where both ILM and RNFL boundaries were reliably segmented and represented measurable RNFLT. Each RNFLT map was resized to 224x224 pixels.

To assess structural validity, we computed several quantitative measures for each RNFLT map: (1) the percentage of invalid tissue, where only eyes with less than 20% invalid tissue were included, and (2) the proportion of valid pixels exceeding 300 µm thickness, where only eyes with this proportion less than 2% were included.^23^ This filtering process removed 1,905 of the original 16,936 samples, leaving 15,031 valid samples for analysis.

### Quality Control and Artifact Correction with EyeLearn

A substantial proportion of OCT-derived RNFLT maps still contained minor or moderate artifacts after the filtering process, including regions of invalid or missing tissue and areas of physiologically implausible thickness. These artifacts both reduced the reliability of the RNFLT maps themselves and also degraded supervision quality for the fundus-to-RNFLT prediction model. Since the accuracy of predicted RNFLT maps depended on the quality of the ground-truth images, we implemented an artifact-correction model to improve the reliability of RNFLT maps before training downstream models.

We adopted the EyeLearn artifact correction model,^23^ a partial-convolution U-Net (PConv-UNet) that inferred missing or corrupted regions while preserving valid retinal structure. The model took as input a colormap-encoded RNFLT map and a corresponding binary validity mask, where valid tissue was marked as 1 and artifacts were marked as 0. During inference, EyeLearn predicted a corrected RNFLT map by filling in the invalid regions while preserving original values in valid regions.

To align our preprocessing with EyeLearn, we resized the RNFLT maps to 256x256 pixels to match EyeLearn’s input resolution and converted to RGB colormaps. Following artifact correction, the predicted RGB output was converted back to micron units, resized to 224x224 pixels, and blended with the original RNFLT maps such that valid pixels remained unchanged and only invalid regions were replaced. The artifact correction model was applied to all 15,031 samples.

### RNFLT Map Prediction Model

We trained a deep learning model to predict RNFLT maps directly from color fundus photographs (Figure 1, Stage 1). The goal was to learn a pixel-wise mapping between fundus image and the OCT-derived RNFLT structure, producing quantitative thickness maps that could later be used for glaucoma classification.

**Figure 1.**
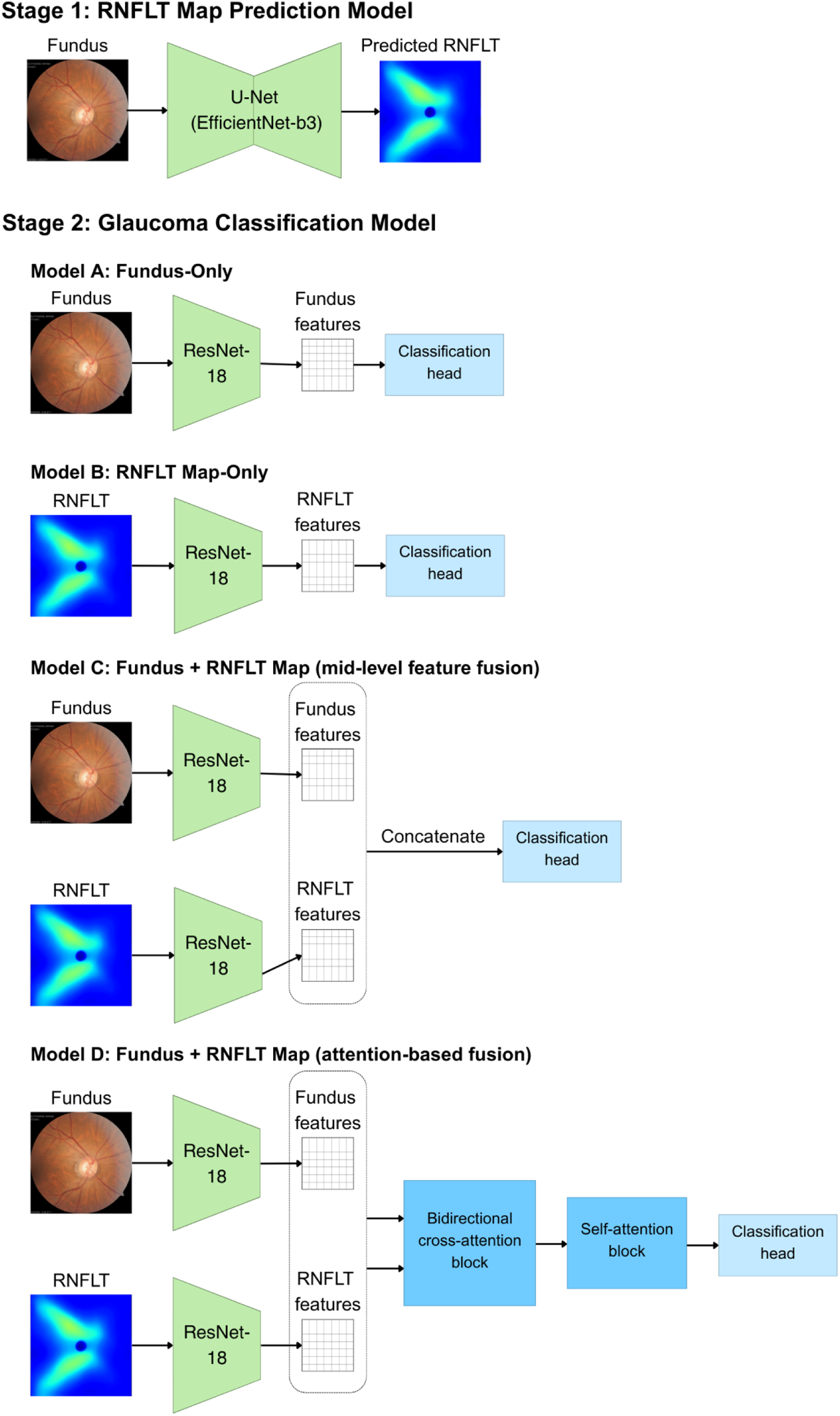
Model architectures in the two-stage pipeline. **Stage 1**: A U-Net-based RNFLT map prediction model that generates OCT-equivalent RNFLT maps from fundus photographs. **Stage 2**: Glaucoma classification models using four input configurations: (Model A) fundus-only model, serving as a baseline for the performance achievable from standard color fundus imaging; (Model B) RNFLT-only model, using either real or predicted RNFLT maps as input; (Model C) mid-level fusion model, which concatenates fundus and RNFLT feature representations before making predictions; and (Model D) attention-based fusion model, which integrates fundus and RNFLT tokens through cross-attention and self-attention to enable deeper multimodal interaction.

The model followed a U-Net architecture^24^ with an encoder-decoder structure. The baseline model used a ResNet-34^25^ encoder pretrained on ImageNet. Alternative encoders (ResNet-50,^25^ EfficientNet-B0,^26^ and EfficientNet-B3^26^) were also evaluated to examine the effect of encoder capacity on performance. The model accepted a 3x224x224 input corresponding to the three RGB channels of the color fundus photograph and produced a 1x224x224 RNFLT map in microns. Skip connections between encoder and decoder layers preserved spatial information.

The model was trained on the filtered and artifact-corrected dataset of 15,031 OCT-fundus pairs. The data were divided into training, validation, and test sets using an 80:10:10 ratio, and samples from the same patient were restricted to a single set to prevent data leakage. Fundus inputs were normalized using ImageNet channel statistics to standardize color and illumination variations across images, while RNFLT maps were standardized by z-score normalization over valid pixels to place thickness values on a consistent numerical scale across samples and reduce the influence of absolute scale differences across eyes.

To better leverage artifact-corrected RNFLT maps produced by the EyeLearn model, we optimized a weighted mean absolute error (MAE) loss between the predicted RNFLT map and the artifact-corrected RNFLT map derived from spectral-domain OCT, which served as the reference standard. MAE presented the average absolute difference between predicted and reference values and provided an interpretable measure of pixel-wise RNFLT disagreement.^27^ The model processed both the corrected RNFLT map and a per-pixel weight map derived from the original tissue mask: the weight was set to 1.0 for pixels that were valid in the original RNFLT map, 0.3 for newly supervised pixels inferred by EyeLearn correction, and 0 for disc and cup pixels, which were excluded from supervision. This approach down-weighted the influence of uncertain regions while still providing weak supervision across previously unsupervised holes, improving overall map continuity.

### Glaucoma Diagnosis

After generating RNFLT maps, we developed a series of models to classify eyes as glaucomatous or non-glaucomatous across different input modalities. These experiments were designed to evaluate the diagnostic value of fundus photographs, RNFLT maps (real or predicted), and their combination through multimodal fusion. ResNet-18 was selected as the backbone for all classifiers due to its lightweight architecture, which offered stable performance for glaucoma classification while reducing overfitting risk.^25^ Using a consistent backbone ensured that performance differences reflected the input modality rather than model complexity.

We first established three single-modality classifiers to assess performance using individual imaging modalities:

1. Fundus-only model: The model received 3-channel RGB fundus photographs as inputs. We applied light augmentation (random flips/rotations and mild brightness/contrast jitter) and trained the model using standard binary cross-entropy loss. This served as a baseline for the performance achievable from standard color fundus imaging (Figure 1, Stage 1: Model A).
2. Real RNFLT-only model: The ResNet-18 was adapted for single-channel RNFLT map input by collapsing the pretrained 3-channel weights. RNFLT inputs were z-score normalized over valid tissue (Figure 1, Stage 2: Model B).
3. Predicted RNFLT-only model: This configuration was identical to the real RNFLT-only model but used the predicted RNFLT maps generated by the fundus-to-RNFLT prediction model described earlier. This enabled direct comparison between models trained on real versus predicted thickness maps (Figure 1, Stage 2: Model B).

To integrate complementary features from fundus photographs and RNFLT maps, we developed multimodal fusion models using a dual-branch mid-level fusion and an attention-based fusion (Figure 1, Stage 2: Model C and D, respectively). The mid-level fusion architecture consisted of two parallel ResNet-18 encoders: one for the 3-channel fundus image and one for the 1-channel RNFLT map.

Feature vectors from the final global average pooling layer of each encoder were concatenated and passed through a fully connected classification head (two linear layers with ReLU activation and dropout). Geometric data augmentations (flips/rotations) were applied identically to both modalities to preserve spatial correspondence, and brightness/contrast jitter was applied only to the fundus. While our primary focus was on predicted RNFLT maps for the fusion model, we also trained a fused fundus and real RNFLT model to establish an approximate upper bound on performance.

This mid-level fusion approach was chosen because it allowed each encoder to learn modality-specific representations before combining them, which has been shown in prior studies to outperform early or late fusion.^28–31^

To further enhance cross-modal feature interaction, we developed an attention-based fusion module that extended the dual-branch architecture with a transformer-style attention mechanism.^32–33^ As illustrated in Figure 1, Stage 2: Model D, the fundus and RNFLT branches each produced a 7x7 spatial feature map from their final convolutional layers. These maps were flattened into tokens, projected into a shared embedding dimension, and supplemented with learned modality embeddings and positional encodings. A [CLS] token was prepended to represent the final classification result, yielding a joint sequence of 99 tokens (1 [CLS] + 49 fundus + 49 RNFLT).

Within this sequence, a transformer performed self-attention and bidirectional cross-attention, allowing the network to dynamically re-weight fundus and RNFLT features based on their relative informativeness. The [CLS] representation was then passed through a normalization layer and an MLP classification head to produce the final glaucoma probability. This attention-based approach enabled adaptive feature weighting between modalities, prioritizing RNFLT-derived structural cues when thickness information was reliable and emphasizing fundus features when the RNFLT signal was less certain. This design maintained architectural consistency with the mid-level fusion baseline, enabling direct performance comparison.

All classifiers were implemented in PyTorch with a ResNet-18 backbone initialized with ImageNet weights. Binary cross-entropy loss was used for optimization, and all models were trained with the Adam optimizer with early stopping based on validation AUC. All experiments used patient-level splits, assigning all images from the same patient to a single set (training, validation, or testing) to prevent information leakage across partitions.

### Evaluation Metrics and Statistical Analysis

We evaluated model performance using the area under the receiver operating characteristic curve (AUC) as the primary metric and reported accuracy descriptively.

To assess statistical significance in model comparisons, we used the DeLong test to compare AUCs between correlated ROC curves. The same test was applied to stratified analyses by demographic groups (race, ethnicity, sex, and age). For glaucoma severity subgroups, AUC could not be computed due to class homogeneity within each subgroup (e.g., all eyes in the “moderate” group have glaucoma). Since AUC measures how well a model distinguishes between two classes and is not meaningful when only one class is present, we used accuracy to evaluate model performance within each severity group.

## Results

### Population Characteristics

The final dataset consisted of 15,031 samples, divided into training (n = 11,882), validation (n = 1,602), and test (n = 1,547) sets. Glaucoma prevalence was 9,231 out of 15,031 (61.4%), and the mean age of patients was 63.8 ± 14.4 years. Among glaucoma cases, the distribution of disease severity was relatively balanced overall, with 21.5% mild, 22.0% moderate, and 17.9% severe (**Table 1**). Patients were primarily White (63.2%), non-Hispanic (86.8%), and slightly more female (53.5%) than male. The largest age group was 60–69 years (29.6%), followed by 70-79 years (25.6%) and 50-59 years (17.0%).

**Table 1.**
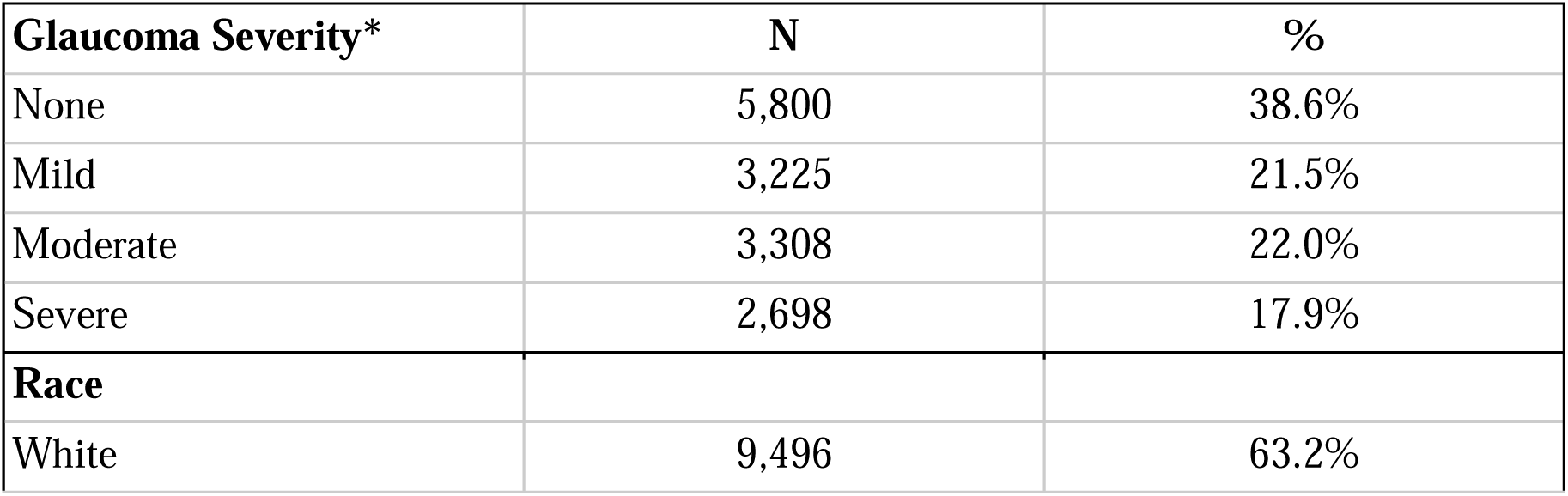
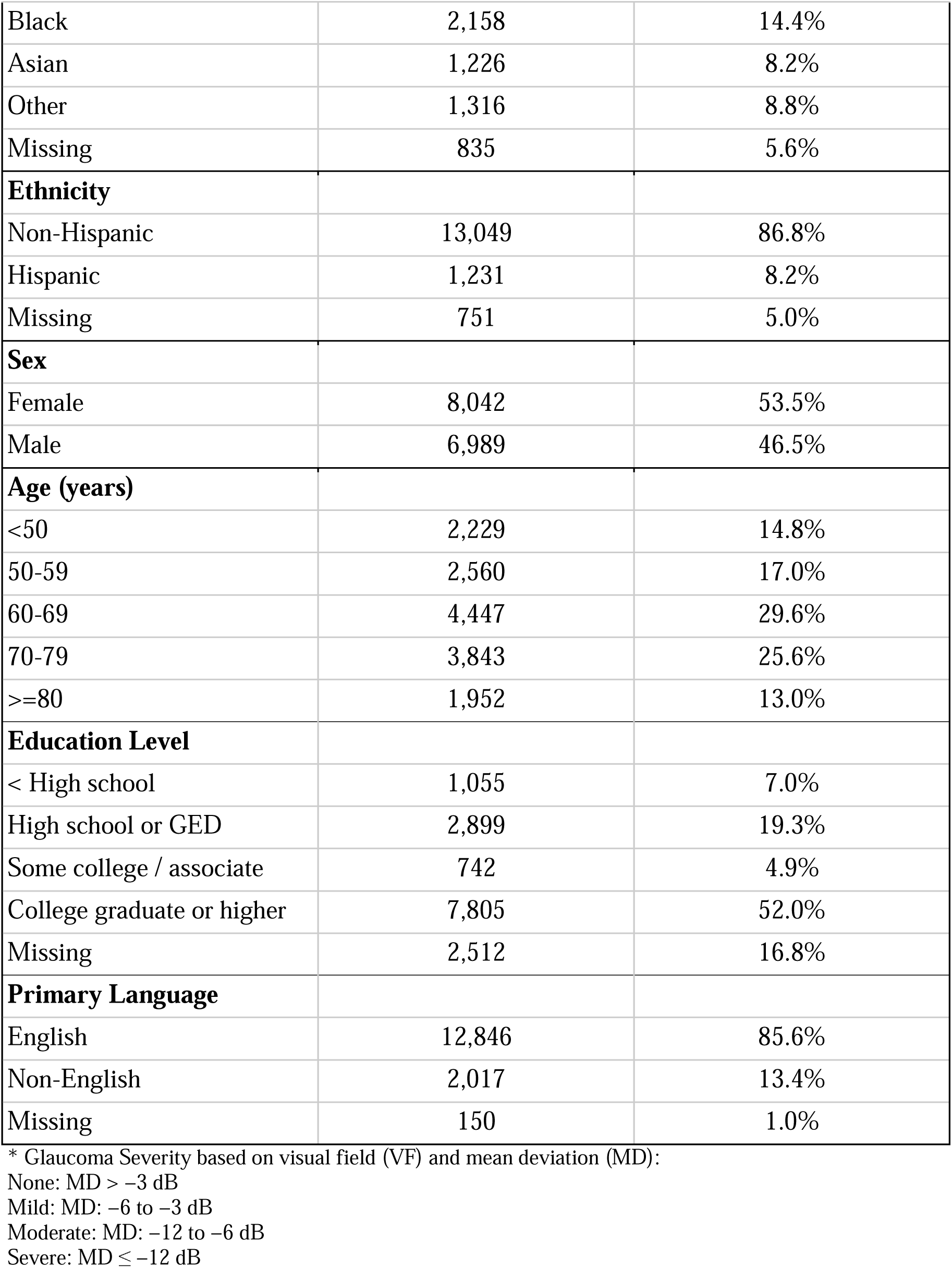
Dataset Distribution by Glaucoma Severity, Demographics, and Social Factors.

| **Glaucoma Severity\*** | **N** | **%** |
| --- | --- | --- |
| None | 5,800 | 38.6% |
| Mild | 3,225 | 21.5% |
| Moderate | 3,308 | 22.0% |
| Severe | 2,698 | 17.9% |
| **Race** |  |  |
| White | 9,496 | 63.2% |
| Black | 2,158 | 14.4% |
| Asian | 1,226 | 8.2% |
| Other | 1,316 | 8.8% |
| Missing | 835 | 5.6% |
| **Ethnicity** |  |  |
| Non-Hispanic | 13,049 | 86.8% |
| Hispanic | 1,231 | 8.2% |
| Missing | 751 | 5.0% |
| **Sex** |  |  |
| Female | 8,042 | 53.5% |
| Male | 6,989 | 46.5% |
| **Age (years)** |  |  |
| <50 | 2,229 | 14.8% |
| 50-59 | 2,560 | 17.0% |
| 60-69 | 4,447 | 29.6% |
| 70-79 | 3,843 | 25.6% |
| >=80 | 1,952 | 13.0% |
| **Education Level** |  |  |
| < High school | 1,055 | 7.0% |
| High school or GED | 2,899 | 19.3% |
| Some college / associate | 742 | 4.9% |
| College graduate or higher | 7,805 | 52.0% |
| Missing | 2,512 | 16.8% |
| **Primary Language** |  |  |
| English | 12,846 | 85.6% |
| Non-English | 2,017 | 13.4% |
| Missing | 150 | 1.0% |

\* Glaucoma Severity based on visual field (VF) and mean deviation (MD):
None: MD > -3 dB
Mild: MD: -6 to -3 dB
Moderate: MD: -12 to -6 dB
Severe: MD ≤ -12 dB

The distributions of race, ethnicity, and sex were similar between the training and testing sets (p > 0.5), while glaucoma severity (p = 0.044) and age (p < 0.001) differed significantly. When all three splits were compared (training, validation, and testing), significant differences were observed in glaucoma severity, race, ethnicity, and age, driven largely by the validation set. These differences may reflect natural variation across patients, as the dataset was split at the patient level to prevent information leakage. Details are provided in Supplementary Table S1.

### RNFLT Map Prediction Performance

We evaluated several encoder backbones for fundus-to-RNFLT prediction, including EfficientNet-B3, EfficientNet-B0, ResNet-34, and ResNet-50. EfficientNet-B3 achieved the strongest performance on the held-out test set with MAE = 15.4 μm and structural similarity index (SSIM) = 0.65, computed over valid tissue (**Table 2**). As discussed earlier, we performed our experiments using the filtered and artifact-corrected dataset. For comparison, a model trained based on the original 16,936 samples before filtering and EyeLearn correction performed worse, with MAE = 24.6 μm and SSIM = 0.59. These results suggest that filtering and artifact correction improved image quality, leading to more reliable structural predictions.

**Table 2.** RNFLT Map Prediction Performance Based on Different Encoder Backbones.

| **Encoder** | **MAE ($\mu\text{m}$)** | **SSIM** |
| --- | --- | --- |
| EfficientNet-B3 | 15.4 | 0.65 |
| EfficientNet-B0 | 16.8 | 0.63 |
| ResNet-34 | 17.3 | 0.60 |
| ResNet-50 | 17.4 | 0.59 |

MAE = mean absolute error; SSIM = structural similarity index.

Figures 2 and 3 illustrate the effect of artifact correction and the quality of the predicted maps. The artifact-correction method, which operated on each OCT scan independently, produced corrected RNFLT maps (Figure 2) with improved continuity in missing or corrupted regions, providing cleaner structural targets for model training. The predicted RNFLT maps (Figure 3) followed the spatial patterns of the artifact-corrected ground truth, capturing major thickness gradients and localized RNFL loss.

**Figure 2.**
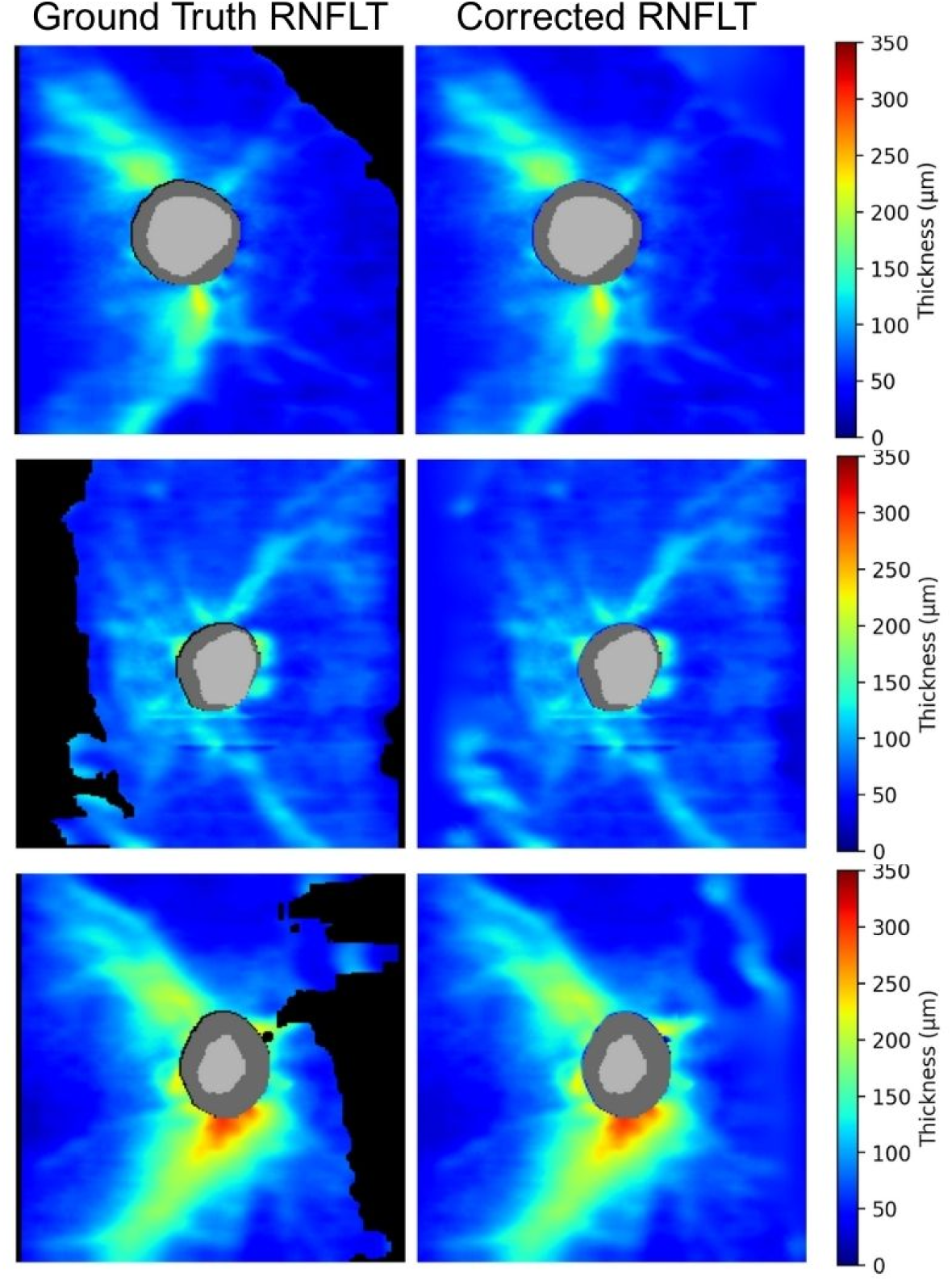
Examples of original OCT-derived RNFLT maps and corresponding artifact-corrected RNFLT maps.

**Figure 3.**
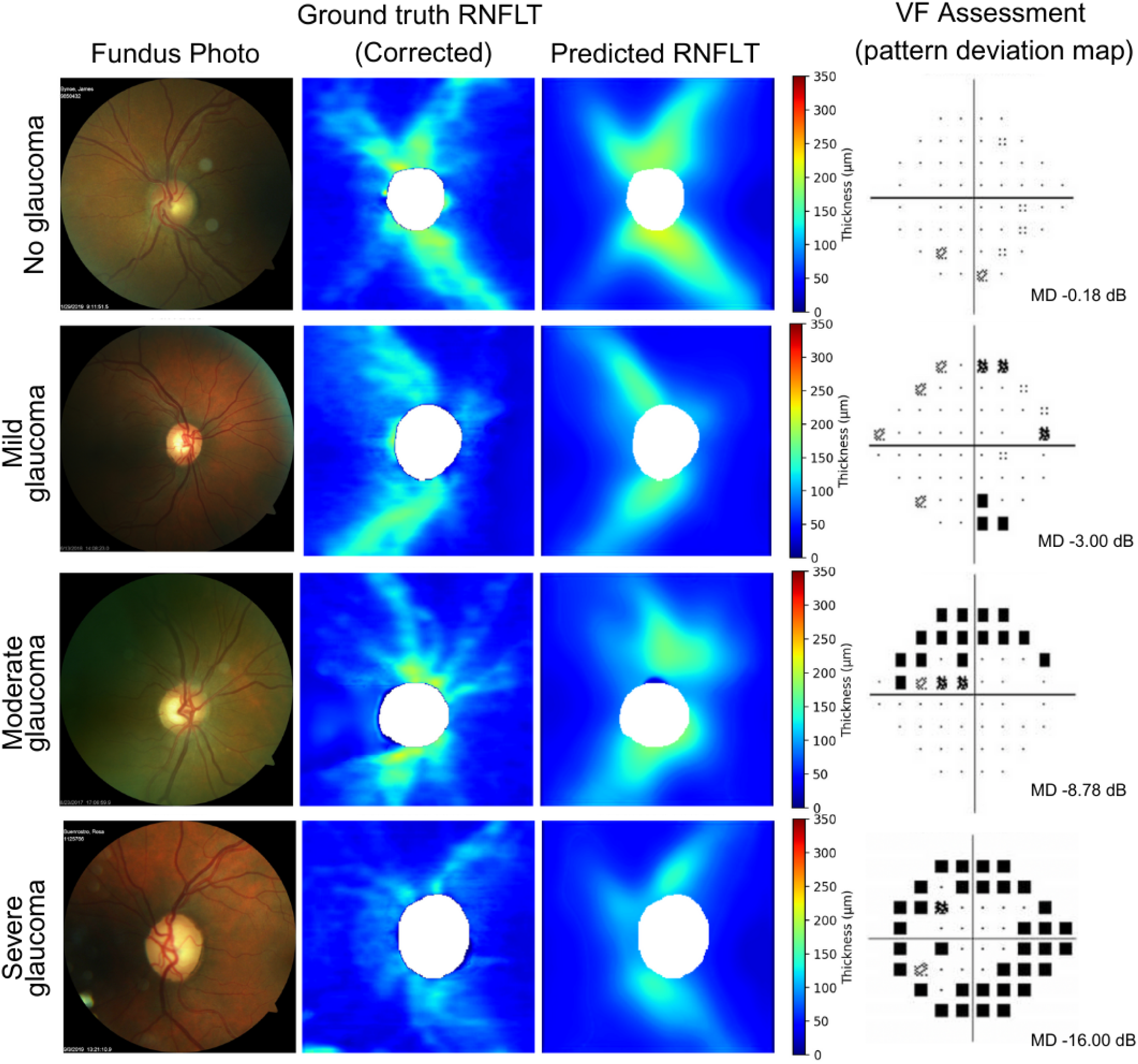
Examples of original fundus photographs, artifact-corrected OCT-derived RNFLT maps, and corresponding predicted RNFLT maps for eyes across glaucoma severity levels (none, mild, moderate, severe glaucoma). Each example includes the Humphrey visual field pattern deviation plot and mean deviation (MD).

### Glaucoma Classification Performance

Table 3 and Figure 4 report the glaucoma diagnostic performance, comparing single-modality models (fundus-only, real-RNFLT-only, predicted-RNFLT-only) and multimodal fusion (fundus + RNFLT). Among the single-modality models, the real RNFLT-only model achieved the highest performance (AUC = 0.903, Accuracy = 83.1%), followed by the predicted RNFLT-only model (AUC = 0.889, Accuracy = 82.0%), both outperforming the fundus-only model (AUC = 0.883, Accuracy = 78.0%) with p values < 0.005 based on the DeLong test. The real RNFLT-only model performed better than the predicted RNFLT-only model (AUC 0.903 vs 0.889, p < 0.005). Both multimodal fusion models (fundus + real RNFLT and fundus + predicted RNFLT) further improved over their single-modality counterparts, with the fusion of fundus and predicted RNFLT achieving an AUC of 0.909 and an accuracy of 82.5%.

**Figure 4.**
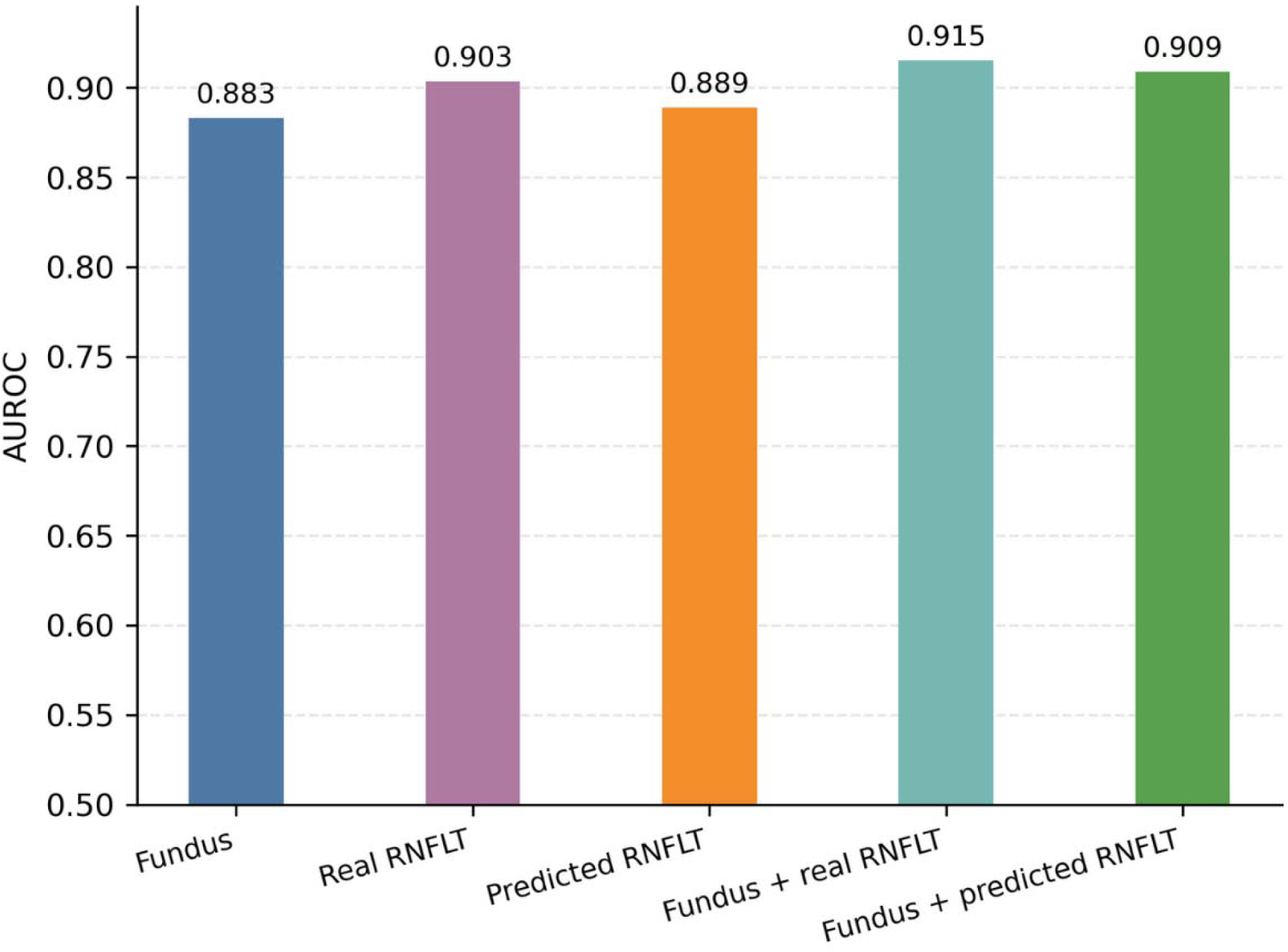
AUCs for glaucoma detection across five input configurations: fundus only, real RNFLT only, predicted RNFLT only, fused fundus + real RNFLT, and fused fundus + predicted RNFLT. Multimodal models are based on attention-based fusion. P values for AUC comparisons are based on the DeLong test, as reported in Table 3. All comparisons were statistically significant (p < 0.005).

**Table 3.** Glaucoma Detection Performance.

|  | AUC | Accuracy | P-value (vs. other models)* |
| --- | --- | --- | --- |
| **Single-Modality** |  |  |  |
| (1) Fundus only | 0.883 | 78.0% |  |
| (2) Real RNFLT Map | 0.903 | 83.1% | vs. (1): <0.005 |
| (3) Predicted RNFLT Map | 0.889 | 82.0% | vs. (1): <0.005<br>vs. (2): <0.005 |
| **Multimodal: Mid-Level Fusion** |  |  |  |
| (4) Fundus + Real RNFLT Map | 0.913 | 84.0% | vs. (1): p < 0.005<br>vs. (2): p < 0.005<br>vs. (3): p < 0.005 |
| (5) Fundus + Predicted RNFLT Map | 0.900 | 81.9% | vs. (1): p < 0.005<br>vs. (2): p < 0.005<br>vs. (3): p < 0.005 |
| **Multimodal: Attention-Based Fusion** |  |  |  |
| (6) Fundus + Real RNFLT Map | 0.915 | 84.2% | vs. (1): p < 0.005<br>vs. (2): p < 0.005<br>vs. (3): p < 0.005 |
| (7) Fundus + Predicted RNFLT Map | 0.909 | 82.5% | vs. (1): p < 0.005<br>vs. (2): p < 0.005<br>vs. (3): p < 0.005 |

\*P-values are based on DeLong tests comparing AUCs between models.

### Stratified Analyses

We further evaluated model performance across disease severity and demographic subgroups, comparing the fundus-only model, the predicted-RNFLT-only model, and the fundus + predicted RNFLT (mid-level fusion) model. Stratified results showed that models incorporating predicted RNFLT maps, either alone or fused with fundus images, yielded higher accuracy or AUC across all categories (Tables 4-8).

**Table 4.** Comparison of Fundus-Only, Predicted-RNFLT-Only, and Fundus + Predicted RNFLT Fusion Models: by Glaucoma Severity.

| **Glaucoma Severity\*** | **N** | **Accuracy** |  |  |
| --- | --- | --- | --- | --- |
|  |  | **(1)**<br>**Fundus** | **(2)**<br>**Predicted RNFLT** | **(3)**<br>**Fused fundus +**<br>**predicted RNFLT** |
| Severe | 2698 (17.9%) | 90.0% | 95.0% | 94.6% |
| Moderate | 3308 (22.0%) | 71.5% | 79.9% | 80.4% |
| Mild | 3225 (21.5%) | 55.3% | 68.1% | 64.0% |
| None | 5800 (38.6%) | 89.0% | 85.7% | 87.1% |

\* Glaucoma Severity based on visual field (VF) and mean deviation (MD):
None: MD > -3 dB
Mild: MD: -6 to -3 dB
Moderate: MD: -12 to -6 dB
Severe: MD ≤ -12 dB
All pairwise comparisons of accuracy between the fundus-only model and the predicted-RNFLT-only and fusion models were statistically significant ( $p < 0.005$ ), based on bootstrapping and paired t-tests.

**Table 5.** Comparison of Fundus-Only, Predicted-RNFLT-Only, and Fundus + Predicted RNFLT Fusion Models: by Race.

| **Race** | **N** | **AUC** |  |  |
| --- | --- | --- | --- | --- |
|  |  | **(1)**<br>**Fundus** | **(2)**<br>**Predicted RNFLT** | **(3)**<br>**Fused fundus +**<br>**predicted RNFLT** |
| White | 9496 (63.2%) | 0.892 | 0.888 | 0.902 |
| Black | 2158 (14.4%) | 0.867 | 0.888 | 0.898 |
| Asian | 1226 (8.2%) | 0.764 | 0.847 | 0.781 |
| Other | 1316 (8.8%) | 0.847 | 0.875 | 0.927 |

All pairwise comparisons of AUC between the fundus-only model and the predicted-RNFLT-only and fusion models were statistically significant ( $p < 0.005$ , DeLong test).

**Table 6.** Comparison of Fundus-Only, Predicted-RNFLT-Only, and Fundus + Predicted RNFLT Fusion Models: by Ethnicity.

| **Ethnicity** | **N** | **AUC** |  |  |
| --- | --- | --- | --- | --- |
|  |  | **(1)**<br>**Fundus** | **(2)**<br>**Predicted RNFLT** | **(3)**<br>**Fused fundus +**<br>**predicted RNFLT** |
| Non-Hispanic | 13049 (86.8%) | 0.895 | 0.893 | 0.905 |
| Hispanic | 1231 (8.2%) | 0.858 | 0.824 | 0.886 |

All pairwise comparisons of AUC between the fundus-only model and the predicted-RNFLT-only and fusion models were statistically significant ( $p < 0.005$ , DeLong test).

**Table 7.** Comparison of Fundus-Only, Predicted-RNFLT-Only, and Fundus + Predicted RNFLT Fusion Models: by Sex.

| **Sex** | **N** | **AUC** |  |  |
| --- | --- | --- | --- | --- |
|  |  | **(1)**<br>**Fundus** | **(2)**<br>**Predicted RNFLT** | **(3)**<br>**Fused fundus +**<br>**predicted RNFLT** |
| Female | 8042 (53.5%) | 0.889 | 0.891 | 0.897 |
| Male | 6989 (46.5%) | 0.879 | 0.889 | 0.904 |

All pairwise comparisons of AUC between the fundus-only model and the predicted-RNFLT-only and fusion models were statistically significant ( $p < 0.005$ , DeLong test).

**Table 8.**
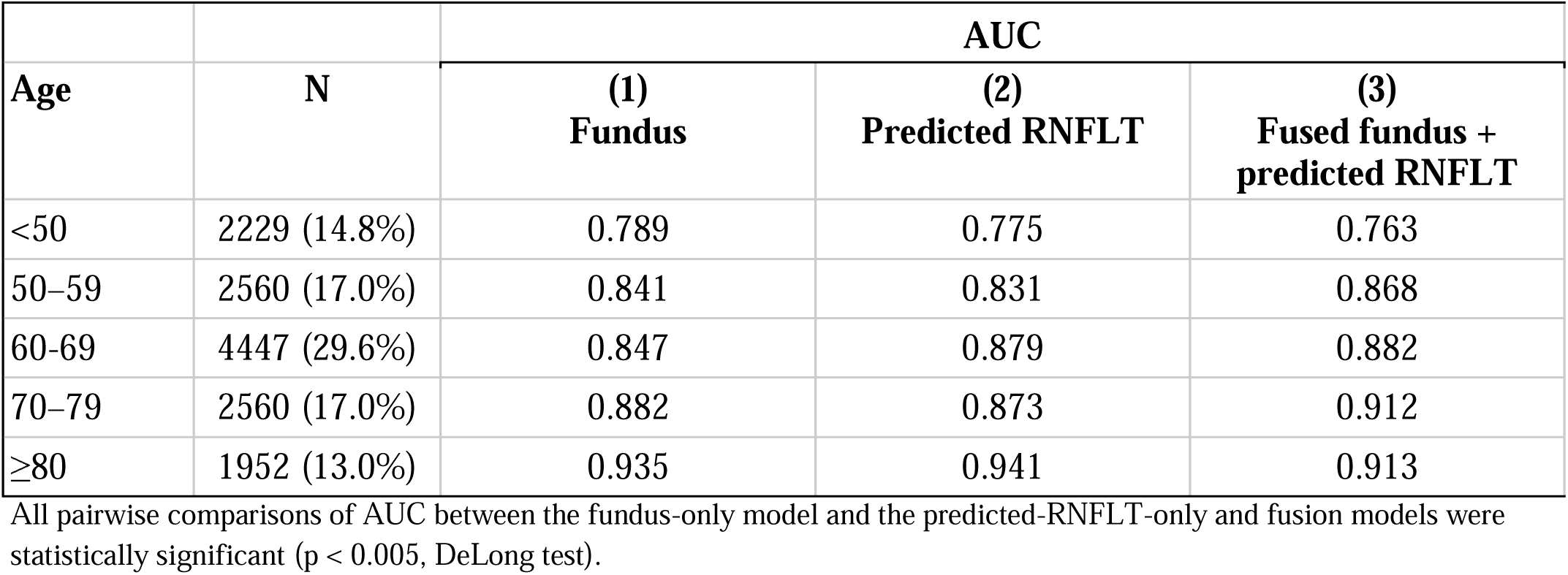
Comparison of Fundus-Only, Predicted-RNFLT-Only, and Fundus + Predicted RNFLT Fusion Models: by Age.

| Age | N | AUC |  |  |
| --- | --- | --- | --- | --- |
|  |  | (1)<br>Fundus | (2)<br>Predicted RNFLT | (3)<br>Fused fundus +<br>predicted RNFLT |
| <50 | 2229 (14.8%) | 0.789 | 0.775 | 0.763 |
| 50–59 | 2560 (17.0%) | 0.841 | 0.831 | 0.868 |
| 60–69 | 4447 (29.6%) | 0.847 | 0.879 | 0.882 |
| 70–79 | 2560 (17.0%) | 0.882 | 0.873 | 0.912 |
| ≥80 | 1952 (13.0%) | 0.935 | 0.941 | 0.913 |

All pairwise comparisons of AUC between the fundus-only model and the predicted-RNFLT-only and fusion models were statistically significant ( $p < 0.005$ , DeLong test).

Across glaucoma severity levels, models using predicted RNFLT maps alone consistently outperformed the fundus-only baseline, particularly for mild and moderate glaucoma, where accuracy increased by 12.8% (p < 0.005) and by 8.4% (p < 0.005), respectively. In contrast, severe cases showed a smaller gain of 5.0% (p < 0.005). Similar patterns were observed for the fused fundus + predicted RNFLT model.

Stratified analyses by demographics showed that the fused fundus + predicted RNFLT model outperformed the fundus-only model, yielding higher AUCs across race, ethnicity, sex, and age. Gains were greater for Black participants (AUC increase of 0.031 from fundus-only to the fusion model) compared with Asian (0.017) and White (0.010) participants, and for Hispanic participants (AUC increase of 0.028) compared with non-Hispanic participants (0.010), suggesting disproportionate benefits in groups that are underrepresented in the dataset. AUC improvements were also larger for males (AUC increase of 0.025) than for females (0.008), and for the 60–69 group (AUC increase of 0.035) relative to other age ranges.

## Discussion

This study demonstrates that OCT–based RNFLT maps can be predicted from fundus photographs with sufficient accuracy to improve glaucoma detection beyond what is achievable using fundus images alone. Among single-modality diagnostic models, predicted RNFLT maps (AUC = 0.889, Accuracy = 82.0%) achieved higher performance than fundus-only models (AUC = 0.883, Accuracy = 78.0%), confirming that the three-dimensional structural information inferred from fundus images captured disease-relevant features beyond the colored photograph. Real RNFLT maps provided the strongest single-modality performance (AUC = 0.903, Accuracy = 83.1%), and the close performance of predicted RNFLT maps demonstrates that synthetic representations derived from fundus photographs can effectively approximate OCT-based structural information. Combining predicted RNFLT maps with fundus photographs through multimodal fusion further improved diagnostic performance, particularly with the attention-based model (AUC = 0.909, Accuracy = 82.5%). These results support that RNFLT-based representations contain structural thickness information that is incremental to the visual cues available in fundus images and that integrating these modalities provides a more comprehensive representation of glaucomatous damage than either input alone.

Between the two fusion models, the attention-based fusion achieved stronger results, indicating that adaptive weighting of informative features enhanced model discriminability. Notably, the attention-based fusion of fundus and predicted RNFLT maps achieved an AUC greater than not only the fundus-only baseline (AUC = 0.909 vs. 0.883, p < 0.005), but also the real-RNFLT-only (AUC = 0.903, p < 0.005) and predicted-RNFLT-only (AUC = 0.889, p < 0.005) models, suggesting that integrating predicted RNFLT maps provided additional structural information that was clinically useful for glaucoma detection.

Models that leveraged predicted RNFLT maps from fundus photographs consistently outperformed fundus-only models even though both used the same source image. This improvement arises because the transformation into an RNFLT representation provided a fundamentally different encoding of structural information. Fundus images contain indirect cues of RNFL health, such as optic disc shape and neuroretinal rim configuration, but these signals may be subtle and difficult to detect, and can be overshadowed by color and lighting variability. The RNFLT prediction model reprojected these cues into a quantitative thickness space that mirrored the structure of true OCT RNFLT maps. As a result, the predicted map acted as a distilled structural representation that more directly encoded features relevant to glaucomatous change.

Using predicted RNFLT maps enhanced diagnostic performance because these maps provided a structured representation of glaucomatous damage that was easier for the model to interpret. The thickness-based input highlighted disease-relevant patterns and reduced noise from color and illumination. This structured input aligned more closely with established clinical biomarkers of RNFL loss, enabling the model to detect glaucoma more reliably than when trained solely on color fundus photographs. As a result, both the predicted-RNFLT model and the fused multimodal models showed improved diagnostic performance.

Our RNFLT map prediction model achieved an MAE of 15.4 μm. This value is higher than those reported in previous studies that predicted global or regional RNFL thickness values, such as Medeiros et al. (2019), who reported an MAE of 7.39 μm for average RNFL thickness.^15^ Predicting a single global value per eye likely results in lower error, as it smooths over localized variability and is less affected by small regional deviations. In contrast, our model predicted a full pixel-wise RNFL thickness map, where error was computed at each valid pixel, making it a more granular and challenging task. As a result, the higher MAE should be interpreted in the context of the richer spatial information provided by the predicted RNFLT maps. The real RNFLT-only model outperformed the predicted RNFLT-only model (AUC 0.903 vs 0.889, p < 0.005), reflecting the advantage of using RNFLT maps derived directly from OCT. In contrast, the predicted maps were generated from fundus photographs, which lack depth information and may reduce fine structural accuracy. Nevertheless, the predicted RNFLT maps, even with relatively high MAE compared with prior studies predicting global or regional RNFLT values, still carried substantial diagnostic value and outperformed the fundus-only model, suggesting their potential as a scalable alternative when OCT is not available.

Stratified analyses revealed that performance gains were greatest in mild and moderate glaucoma cases, where structural damage is subtle and difficult to discern from fundus photographs. Predicted RNFLT maps enhanced detection in these cases by making subtle nerve fiber thinning more explicit through a spatially structured thickness map. Because early glaucoma often lacks prominent optic disc changes, the ability to highlight regional RNFL loss likely provided an advantage over fundus-only models and improved detection of early and mid-stage disease.

In addition, greater improvements were observed in Black and Asian participants compared with White participants, in Hispanic compared with Non-Hispanic participants, and in the 60–69-year subgroup compared with other age groups. This result may reflect how RNFLT maps helped reduce model reliance on demographic-specific visual patterns, such as pigmentation, brightness, or vessel contrast, that vary in fundus images and can introduce bias. By focusing the model on structural information grounded in OCT-like representations, RNFLT maps likely enabled more consistent performance across diverse populations.

Previous “machine-to-machine” studies have shown that global or regional RNFLT values can be estimated from fundus photographs, but these models did not localize damage or support visual interpretation.^15–19^ Our study advances the field by generating continuous pixel-wise RNFLT maps with artifact-corrected supervision and demonstrating measurable improvement in clinical utility for glaucoma diagnosis. The ability to derive quantitative RNFLT maps from widely available fundus cameras has significant clinical potential. In many primary care and community settings, OCT imaging remains inaccessible. By providing OCT-like structural information from a single fundus image, the proposed approach enables OCT-like structural assessment using existing fundus infrastructure, providing interpretable topographic thickness maps rather than qualitative disc photographs alone.

Because these predicted maps approximate the format and units of true OCT-derived RNFLT, they could be integrated into existing glaucoma evaluation pipelines, facilitating reliable and objective screening.

Furthermore, our multimodal fusion experiments confirmed that combining fundus and predicted RNFLT information improved diagnostic accuracy, consistent with prior evidence that fundus and OCT capture complementary aspects of glaucomatous damage. The observed improvements in mild and moderate disease suggest that this model captured image features indicative of early structural damage more effectively than fundus-only models.

We also observed demographic variability in model performance and found that predicted RNFLT maps and multimodal fusion reduced the magnitude of some differences relative to fundus-only models. These results suggest that incorporating structural information, real or predicted, has the potential to improve diagnostic consistency across diverse populations. Overall, the stratified results suggest that integrating predicted RNFLT maps improved diagnostic performance across disease severity and demographic subgroups, with the greatest benefits observed in cases where structural cues were subtle and in groups that were underrepresented in the dataset.

Several limitations should be acknowledged. First, predicted RNFLT maps were learned from OCT-derived targets and may have inherited residual noise or segmentation artifacts despite artifact-correction procedures. Second, this was a retrospective single-center study conducted at a tertiary eye hospital. Although the dataset was large and well curated, it may differ from screening populations, as the prevalence of glaucoma in this cohort was higher than would typically be expected. External validation is needed to confirm generalizability. Third, the fundus photographs were captured by skilled ophthalmic technicians under controlled clinical conditions and using a single imaging device. Image quality and device variability in primary care or community-based screening settings may be more variable, which could affect model performance. Fourth, although the synthesized RNFLT maps enhanced model interpretability, prospective studies are needed to determine how clinicians interpret and act upon these synthetic outputs in real-world workflows. Fifth, the overall MAE of the predicted RNFLT maps remained relatively high, reflecting the difficulty of inferring fine structural details from fundus photographs alone. Reducing this error through model architecture improvements or the incorporation of additional anatomical features, such as axial length, may improve downstream classification. Sixth, although relative gains were greatest in mild glaucoma cases, absolute performance in this group remains modest, likely due to the challenge of detecting early, subtle structural loss. In addition, our dataset excluded cases with borderline VF damage (mean deviation between -1 dB and -3 dB), which may have limited the representation of early-stage disease where structural changes precede functional loss. As a result, model performance in mild cases was partly affected by the dataset definitions. Future work should investigate methods to improve performance in early disease detection. Finally, demographic subgroup analyses should be interpreted cautiously, given uneven sample sizes.

## Conclusion

We present a pipeline that predicts OCT-grade structural information from fundus photographs and uses it to improve glaucoma detection. Our fundus-to-RNFLT model generates quantitative, topographic thickness maps using artifact-corrected supervision and per-pixel quality weighting, demonstrating that clinically meaningful RNFLT structure can be inferred directly from widely available fundus images. In downstream diagnostic evaluation, RNFLT-based models outperformed fundus photo-only baselines, and multimodal fusion of fundus photos and RNFLT maps yielded the strongest results. Stratified analyses showed the greatest gains in mild and moderate disease and in underrepresented demographic groups, suggesting that synthesized RNFLT maps can help detect subtle damage and has the potential to reduce disparities in model performance. These findings offer a practical pathway to extend OCT-like structural assessment to settings where OCT is unavailable, by leveraging existing fundus cameras to generate interpretable RNFLT maps and enhance diagnostic accuracy.

## Data Availability

The data used in this study are derived from patient records at Massachusetts Eye and Ear and are not publicly available due to privacy restrictions.

## References

1. Tham YC, Li X, Wong TY, Quigley HA, Aung T, Cheng CY. Global prevalence of glaucoma and projections of glaucoma burden through 2040: a systematic review and meta-analysis. Ophthalmology. 2014;121(11):2081–2090. doi:10.1016/j.ophtha.2014.05.013

2. About glaucoma. Vision and Eye Health. https://www.cdc.gov/vision-health/about-eye-disorders/glaucoma.html#:∼:text=Facts%20about%20glaucoma,your%20vision%20health%20is%20key. Published May 15, 2024.

3. Geevarghese A, Wollstein G, Ishikawa H, Schuman JS. Optical coherence tomography and glaucoma. Annu Rev Vis Sci. 2021;7(1):693–726. doi:10.1146/annurev-vision-100419-111350

4. Li A, Thompson AC, Asrani S. Impact of artifacts from optical coherence tomography retinal nerve fiber layer and macula scans on detection of glaucoma progression. Am J Ophthalmol. 2021;221:235–245. doi:10.1016/j.ajo.2020.08.018

5. Beniz LAF, Campos VP, Medeiros FA. Optical coherence tomography versus optic disc photo assessment in glaucoma screening. J Glaucoma. 2024;33(Suppl 1):S21–S25. doi:10.1097/IJG.0000000000002392

6. Grzybowski A, Jin K, Zhou J, et al. Retina fundus photograph-based artificial intelligence algorithms in medicine: A systematic review. Ophthalmol Ther. 2024;13(8):2125–2149. doi:10.1007/s40123-024-00981-4

7. Reus NJ, Lemij HG, Garway-Heath DF, et al. Clinical assessment of stereoscopic optic disc photographs for glaucoma: the European Optic Disc Assessment Trial. Ophthalmology. 2010;117(4):717–723. doi:10.1016/j.ophtha.2009.09.026

8. Chauhan BC, McCormick TA, Nicolela MT, LeBlanc RP. Optic disc and visual field changes in a prospective longitudinal study of patients with glaucoma: comparison of scanning laser tomography with conventional perimetry and optic disc photography. Arch Ophthalmol. 2001;119(10):1492–1499. doi:10.1001/archopht.119.10.1492

9. Khan SM, Liu X, Nath S, et al. A global review of publicly available datasets for ophthalmological imaging: barriers to access, usability, and generalisability. Lancet Digit Health. 2021;3(1):e51-e66. doi:10.1016/S2589-7500(20)30240-5

10. Asaoka R, Tanito M, Shibata N, et al. Validation of a deep learning model to screen for glaucoma using images from different fundus cameras and data augmentation. Ophthalmol Glaucoma. 2019;2(4):224–231. doi:10.1016/j.ogla.2019.03.008

11. Li Z, He Y, Keel S, Meng W, Chang RT, He M. Efficacy of a deep learning system for detecting glaucomatous optic neuropathy based on color fundus photographs. Ophthalmology. 2018;125(8):1199–1206. doi:10.1016/j.ophtha.2018.01.02

12. Nguyen V, Iyengar S, Rasheed H, et al. Comparison of deep learning and clinician performance for detecting referable glaucoma from fundus photographs in a safety net population. Ophthalmol Sci. 2025;5(4):100751. doi:10.1016/j.xops.2025.100751

13. Katuru A, Chung IY, Majid I, Shen LQ, Wang M. Deep learning with disc photos or OCT scans in glaucoma detection. Ophthalmol Sci. 2025;5(6):100877. doi:10.1016/j.xops.2025.100877

14. Kim KE, Kim JM, Song JE, Kee C, Han JC, Hyun SH. Development and validation of a deep learning system for diagnosing glaucoma using optical coherence tomography. J Clin Med. 2020;9(7):2167. doi:10.3390/jcm9072167

15. Medeiros FA, Jammal AA, Thompson AC. From machine to machine: An OCT-trained deep learning algorithm for objective quantification of glaucomatous damage in fundus photographs. Ophthalmology. 2019;126(4):513–521. doi:10.1016/j.ophtha.2018.12.033

16. Jammal AA, Thompson AC, Mariottoni EB, et al. Human versus machine: Comparing a deep learning algorithm to human gradings for detecting glaucoma on fundus photographs. Am J Ophthalmol. 2020;211:123–131. doi:10.1016/j.ajo.2019.11.006

17. Yang H, Ahn Y, Askaruly S, You JS, Kim SW, Jung W. Deep learning-based glaucoma screening using regional RNFL thickness in fundus photography. Diagnostics (Basel). 2022;12(11):2894. doi:10.3390/diagnostics12112894

18. Xu Y, Liu H, Sun R, et al. Deep learning for predicting circular retinal nerve fiber layer thickness from fundus photographs and diagnosing glaucoma. Heliyon. 2024;10(13):e33813. doi:10.1016/j.heliyon.2024.e33813

19. Liu JC, Jammal AA, Scherer R, et al. Predicting retinal nerve fiber layer thickness from Ocular Hypertension Treatment Study optic disc photographs. JAMA Ophthalmol. 2025;143(8):652–659. doi:10.1001/jamaophthalmol.2025.1740

20. Chen HSL, Chen GA, Syu JY, et al. Early glaucoma detection by using style transfer to predict retinal nerve fiber layer thickness distribution on the fundus photograph. Ophthalmol Sci. 2022;2(3):100180. doi:10.1016/j.xops.2022.100180

21. Hodapp E, Parrish RK, Anderson DR, Hodapp E, Parrish RK, Anderson DR. Book Clinical decisions in glaucoma. In: Mosby; C1993 Library Catalog; MMS ID 997269853406676; ISBN 9780801667992; ISBN 0801667992; NLM Unique.; 1938.

22. Chauhan BC, Garway-Heath DF, Goñi FJ, et al. Practical recommendations for measuring rates of visual field change in glaucoma. Br J Ophthalmol. 2008;92(4):569–573. doi:10.1136/bjo.2007.135012

23. Shi M, Lokhande A, Fazli MS, et al. Artifact-tolerant clustering-guided contrastive embedding learning for ophthalmic images in glaucoma. IEEE J Biomed Health Inform. 2023;27(9):4329–4340. doi:10.1109/JBHI.2023.3288830

24. Ronneberger O, Fischer P, Brox T. U-Net: Convolutional Networks for Biomedical Image Segmentation. In: Lecture Notes in Computer Science. Springer International Publishing; 2015:234-241. doi.org/10.1007/978-3-319-24574-4_28

25. He K, Zhang X, Ren S, Sun J. Deep residual learning for image recognition. In: 2016 IEEE Conference on Computer Vision and Pattern Recognition (CVPR). IEEE; 2016:770-778. doi.org/10.1109/CVPR.2016.90

26. Tan M, Le QV. EfficientNet: Rethinking model scaling for convolutional Neural Networks. Chaudhuri K, Salakhutdinov R, eds. *arXiv [csLG]*. Published online 09--15 Jun 2019:6105-6114. https://proceedings.mlr.press/v97/tan19a.html

27. Nouri-Mahdavi K, Mohammadzadeh V, Rabiolo A, Edalati K, Caprioli J, Yousefi S. Prediction of visual field progression from OCT structural measures in moderate to advanced glaucoma. Am J Ophthalmol. 2021;226:172–181. doi:10.1016/j.ajo.2021.01.023

28. Yi S, Zhang G, Qian C, Lu Y, Zhong H, He J. A multimodal classification architecture for the severity diagnosis of glaucoma based on deep learning. Front Neurosci. 2022;16:939472. doi:10.3389/fnins.2022.939472

29. Shehryar T, Akram MU, Khalid S, et al. Improved automated detection of glaucoma by correlating fundus and SD OCT image analysis. Int J Imaging Syst Technol. 2020;30(4):1046–1065. doi:10.1002/ima.22413

30. Yoo TK, Choi JY, Seo JG, Ramasubramanian B, Selvaperumal S, Kim DW. The possibility of the combination of OCT and fundus images for improving the diagnostic accuracy of deep learning for age-related macular degeneration: a preliminary experiment. Med Biol Eng Comput. 2019;57(3):677–687. doi:10.1007/s11517-018-1915-z

31. Islam S, Deo RC, Barua PD, Soar J, Acharya UR. Novel deep learning model for glaucoma detection using fusion of fundus and optical coherence tomography images. Sensors (Basel). 2025;25(14):4337. doi:10.3390/s25144337

32. Huynh J, Gonzalez R, Walker E, et al. Multimodal transformer model to detect glaucoma from OCT and retinal nerve fiber layer (RNFL) thickness. Invest Ophthalmol Vis Sci. 2023;64(8).

33. Gao H, Zhao S, Zheng G, et al. Using a dual-stream attention neural network to characterize mild cognitive impairment based on retinal images. Comput Biol Med. 2023;166(107411):107411. doi:10.1016/j.compbiomed.2023.107411

