## Supplementary Table S1 for "Deriving OCT-Equivalent Retinal Nerve Fiber Layer Thickness Maps from Fundus Photographs with Deep Learning Improves Glaucoma Diagnosis"

Supplementary Table S1. Comparison of Dataset Distribution by Glaucoma Severity and Demographics between Training, Validation, and Testing Sets

|  | **Training**  **(n=11,882)** | **Validation (n=1,602)** | **Testing**  **(n=1,547)** | **P Value**  **(Train vs Test)** | **P Value**  **(Train vs Val**  **vs Test)** |
| --- | --- | --- | --- | --- | --- |
| **Glaucoma Severity** |  |  |  | 0.044 | 0.034 |
| None | 4,593 (38.7%) | 569 (35.5%) | 638 (41.2%) |  |  |
| Mild | 2,533 (21.3%) | 347 (21.7%) | 345 (22.3%) |  |  |
| Moderate | 2,629 (22.1%) | 356 (22.2%) | 323 (20.9%) |  |  |
| Severe | 2,127 (17.9%) | 330 (20.6%) | 241 (15.6%) |  |  |
| **Race** |  |  |  | 0.531 | < .001 |
| White | 7,574 (63.7%) | 969 (60.5%) | 953 (61.6%) |  |  |
| Black | 1,701 (14.3%) | 222 (13.9%) | 235 (15.2%) |  |  |
| Asian | 982 (8.3%) | 108 (6.7%) | 136 (8.8%) |  |  |
| Other | 1,035 (8.7%) | 144 (9.0%) | 137 (8.9%) |  |  |
| Missing | 590 (5.0%) | 159 (9.9%) | 86 (5.6%) |  |  |
| **Ethnicity** |  |  |  | 0.849 | < .001 |
| Non-Hispanic | 10,377 (87.3%) | 1,319 (82.3%) | 1,353 (87.5%) |  |  |
| Hispanic | 977 (8.2%) | 151 (9.4%) | 122 (7.9%) |  |  |
| Missing | 528 (4.4%) | 151 (9.4%) | 72 (4.7%) |  |  |
| **Sex** |  |  |  | 0.543 | 0.762 |
| Female | 6,339 (53.3%) | 865 (54.0%) | 838 (54.2%) |  |  |
| Male | 5,543 (46.7%) | 737 (46.0%) | 709 (45.8%) |  |  |
| **Age (years)** |  |  |  | < .001 | < .001 |
| <50 | 1,833 (15.4%) | 207 (12.9%) | 189 (12.2%) |  |  |
| 50-59 | 2,078 (17.5%) | 248 (15.5%) | 234 (15.1%) |  |  |
| 60-69 | 3,446 (29.0%) | 512 (32.0%) | 489 (31.6%) |  |  |
| 70-79 | 3,027 (25.5%) | 405 (25.3%) | 411 (26.6%) |  |  |
| >=80 | 1,498 (12.6%) | 230 (14.4%) | 224 (14.5%) |  |  |

Glaucoma severity was defined based on visual field (VF) mean deviation (MD): none (MD > −3 dB), mild (−6 to −3 dB), moderate (−12 to −6 dB), and severe (≤ −12 dB). dB = decibels.
